# The projected cost-effectiveness and budget impact of Alternative HPV Vaccines in Senegal: A Modelling Study

**DOI:** 10.1101/2025.10.08.25337573

**Authors:** Oumaima Laraj, Maryam Diarra, Cheikh Talla, Rokhaya Diop, Ousseynou Badiane, Karell Pelle, Amira Kebir, Cheikh Loucoubar, Slimane Ben Miled

## Abstract

**Background:** Cervical cancer remains a leading cause of morbidity and mortality among women in Senegal. In 2018, with support from Gavi, Senegal introduced HPV vaccination into its national immunization program using a two-dose Gardasil-4 schedule for 9-year-old girls. As Senegal approaches transition from Gavi support, full procurement costs will shift to the government, necessitating evidence-based decisions about the most cost-effective vaccination strategies going forward. This study evaluates the health and economic value of alternative HPV vaccination options to inform sustainable and equitable national policy.

**Methods:** We used a static cohort model to assess the cost-effectiveness and budget impact of 23 HPV vaccination strategies varying by vaccine type (Gardasil-4, Gardasil-9, Cervarix, Cecolin), dose number (one vs. two), and coverage level (current, 70%, WHO target). The analysis simulates health and cost outcomes for 9-year-old girls vaccinated annually from 2019 to 2035. Outcomes included cervical cancer cases, deaths, and disability-adjusted life years (DALYs) averted. Costs were evaluated from a government perspective in 2023 USD, with a 3% discount rate applied. Input parameters were derived from GLOBOCAN 2020, peer-reviewed literature, and national data. Sensitivity analyses explored uncertainty around key assumptions.

**Results:** All HPV vaccination strategies were cost-effective under a willingness-to-pay threshold of 30% of GDP per capita (USD 450 per DALY averted). Cecolin-based strategies, particularly one-dose schedules with expanded coverage, consistently yielded the most favorable cost-effectiveness ratios. Gardasil-9 offered the greatest absolute health benefits (up to 112,866 DALYs averted) but required significantly higher programmatic investments, limiting its cost-effectiveness unless vaccine prices were reduced. Five strategies mainly involving Cecolin and one-dose Gardasil-9) were efficient under national willingness-to-pay thresholds. At lower thresholds, such as 7% of GDP per capita or USD 100, one-dose Cecolin with 90% coverage was optimal.

**Conclusion:** HPV vaccination remains a highly cost-effective intervention in Senegal across multiple scenarios. Cecolin-based strategies, especially one-dose regimens, offer a scalable and economically attractive approach post-Gavi. Gardasil-9 could be viable with substantial price reductions. As Senegal prepares for Gavi transition, aligning vaccine choice with both cost-effectiveness and implementation feasibility will be essential to ensuring long-term program sustainability and equitable health outcomes.

## 1 Introduction

Cervical cancer is the fourth most common cancer worldwide and remains a major burden in socioeconomically underserved regions of low- and middle-income countries (LMICs), particularly in Sub-Saharan Africa (SSA) [40]. Among the 20 countries with the highest cervical cancer burden, 19 are located in SSA [29]. According to the World Health Organization (WHO), cervical cancer causes more than 300,000 deaths annually and is projected to kill over 443,000 people by 2030, nearly 90% of which will occur in SSA, despite the disease being almost entirely preventable [15]. In Senegal, cervical cancer is the second most common cancer among women and the leading cause of cancer-related mortality, with an age-standardized incidence rate of 34.3 per 100,000 (more than twice the global average) and an estimated 1,327 deaths in 2023 [23].

In response to this challenge, the WHO launched a global strategy in 2018 to eliminate cervical cancer as a public health problem by 2030. The “90-70-90” targets aim to ensure that 90% of girls are fully vaccinated against HPV before the age of 15, that 70% of women aged 35–45 are screened, and that 90% of women diagnosed with cervical cancer receive appropriate treatment [48].

Six prophylactic HPV vaccines are currently licensed worldwide, all produced using recombinant DNA technology. These include three bivalent vaccines (Cervarix, Cecolin, and Walrinvax), two quadrivalent vaccines (Gardasil-4 and Cervavac), and one nonavalent vaccine (Gardasil-9) [10]. While all are internationally available, only Cervarix, Gardasil-4, and Gardasil-9 are currently accessible in SSA. Optimizing HPV vaccination strategies in resource-limited settings is essential to fully realize the benefits of these programs and move toward the goal of cervical cancer elimination [31].

Only 10% of Senegalese women have ever been screened for cervical cancer, but efforts are underway to strengthen and expand screening and treatment services [12]. With Gavi support, Senegal conducted a school-based HPV vaccine demonstration project from 2014 to 2016 in Dakar Ouest and Meckhe districts, achieving 90% administrative coverage among nine-year-old girls, with a total of 11,232 doses administered [7]. Building on this success, HPV vaccination was integrated into the national routine immunization program in late 2018, again with Gavi support, using a two-dose schedule of Gardasil-4 delivered through health facilities, schools, and community outreach [9].

Coverage trends since the national rollout have been uneven. In 2019, 89% of eligible girls received the first dose, but only 25% completed the second. Coverage dropped substantially in 2020, with 47% for the first dose and 32% for the second, and showed only partial recovery in 2021 (65% and 33%, respectively). However, this improvement was not sustained: between 2022 and 2024, coverage declined again and stabilized at around 47% for the first dose and 25% for the second [49]. In our analysis, vaccination coverage is assumed to increase from 47% in 2024 to 60% in 2025, ultimately reaching the WHO target of 90% by 2030.

Globally, HPV genotypes 16 and 18 are most commonly associated with cervical cancer, though the distribution of other high-risk types varies geographically [46]. In Senegal, recent evidence shows that most women with histologically confirmed cervical cancer are co-infected with multiple high-risk HPV types, with genotypes 16, 18, 45, 33, and 59 being the most frequent [13]. Niane et al. [26] highlighted that the nonavalent Gardasil-9 vaccine, which targets nine HPV genotypes (6, 11, 16, 18, 31, 33, 45, 52, and 58), could provide broader protection for adolescent girls in Senegal.

There is increasing evidence that one or two doses of HPV vaccine provide comparable protection [25]. A single-dose regimen could simplify and accelerate implementation, reduce costs, and facilitate higher coverage, particularly in African contexts [36]. In Kenya, both bivalent and nonavalent single-dose vaccines demonstrated high efficacy against persistent oncogenic HPV infections, comparable to multidose schedules among women aged 15–20 years [5]. Economic evaluations in Kenya further showed that single-dose vaccination offers similar health benefits at lower cost [42, 32]. The recently approved single-dose vaccine Cecolin has shown efficacy comparable to Cervarix, Gardasil-4, and Gardasil-9, but at lower cost [3], with a study in Burkina Faso identifying Cecolin as the most cost-effective option [27]. In Senegal, one of the first nationally representative costing studies of HPV vaccine introduction using routine immunization strategies has been conducted [7]. However, although HPV vaccination has been found to be cost-effective in several LMICs, including those in SSA [20, 45], no formal cost-effectiveness analysis has yet been carried out in Senegal.

This study aims to evaluate the potential health and economic impact of Senegal’s national HPV vaccination program, currently based on the quadrivalent Gardasil-4 vaccine. It further explores alternative strategies, including the introduction of Gavi-supported bivalent vaccines (Cecolin and Cervarix), the nonavalent Gardasil-9 vaccine, and a simplified single-dose schedule. By modeling different coverage trajectories, this analysis seeks to inform national decision-makers on how to maximize the effectiveness, efficiency, and sustainability of HPV vaccination efforts in Senegal.

The paper is organized as follows. Section 2 describes the study design, modeling approach, input parameters, and the cost-effectiveness and sensitivity analyses. In section 3, we present the main results, including the cost-effectiveness of HPV vaccination strategies, identification of optimal approaches, the impact of Gardasil-9 price reductions, and sensitivity analyses. Section 4 discusses and interprets the findings in relation to existing literature, highlights policy and programmatic implications for Senegal, acknowledges study limitations, and outlines directions for future research. Finally, we summarize the key insights and their relevance for sustainable HPV vaccination policy.

## 2 Methods

### 2.1 Study design

From a government perspective, we assessed the cost-effectiveness of various HPV vaccination strategies, considering different vaccine products (Cecolin, Cervarix, Gardasil-4, and Gardasil-9), dose schedules, and coverage scenarios. Each strategy was compared not only to the current baseline scenario using Gardasil-4 (with no changes in screening practices) but also to alternative vaccine options. In our base case, we evaluated the potential impact and cost-effectiveness of vaccinating seventeen annual cohorts of 9-year-old girls over a 17-year period (2019– 2035) in Senegal. We considered two dosing strategies: continuing the current two-dose schedule for all four vaccines and transitioning to a single-dose schedule starting in 2025. Furthermore, we analyzed three coverage scenarios: **Current Trend**, reflecting historical uptake from 2019 to 2024; **Moderate Increase**, assuming a gradual improvement in coverage; and **90% Coverage**, aiming to reach 90% first-dose coverage in alignment with WHO targets (Table 1).

**Table 1:**
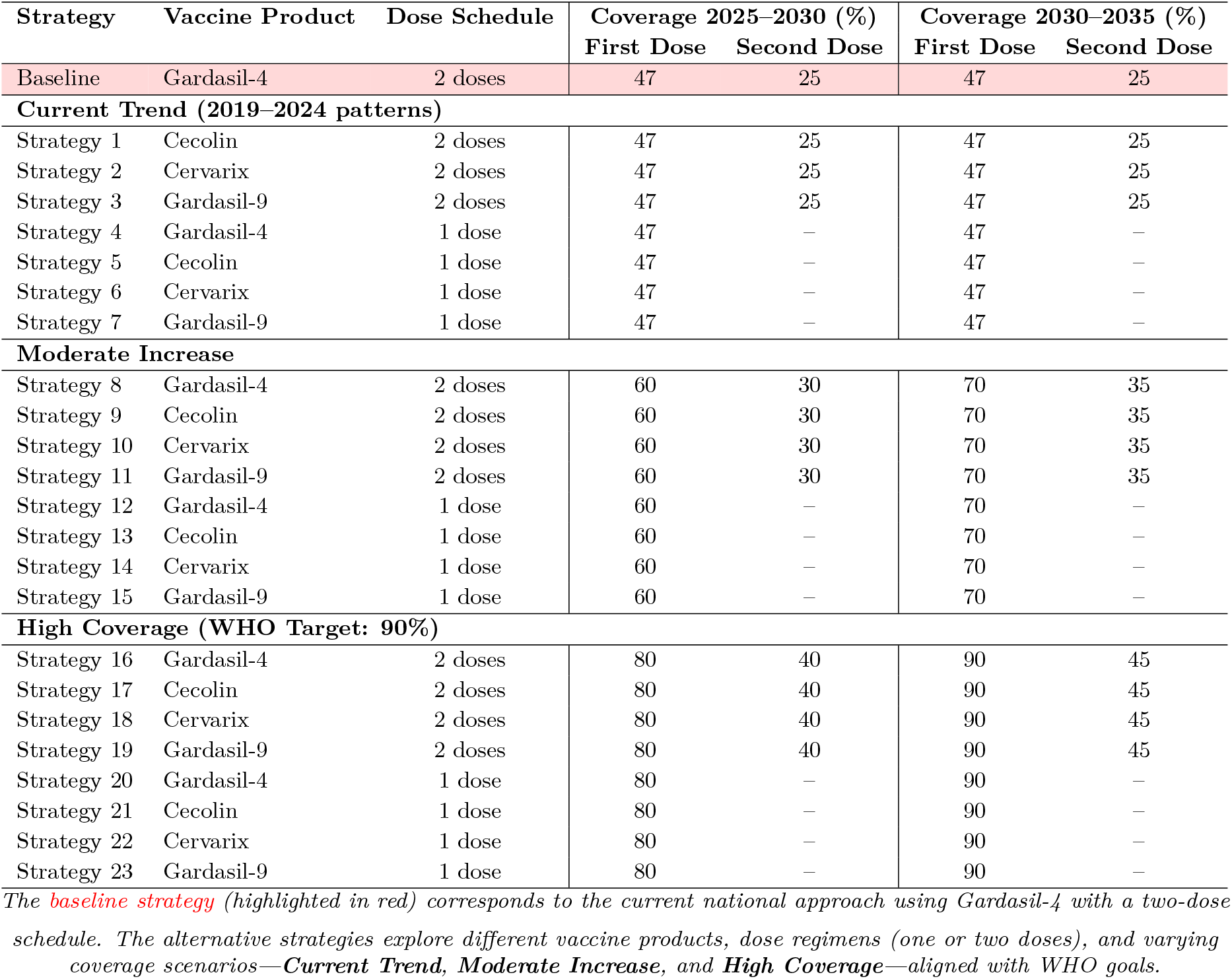
HPV Vaccination Strategies in Senegal (2025–2035).

A multidisciplinary group of experts has been engaged in a series of stakeholder consultation meetings starting from March 2025 to provide feedback on the inputs and assumptions used in the analysis. Participants included the Head of the Immunisation Department of the Senegalese Ministry of Health, members of the Biostatistics and Modeling team at the Institut Pasteur in Dakar, and other key stakeholders.

### 2.2 Modelling approach

We used the UNIVAC tool, an Excel-based static cohort model that estimates proportionate outcomes [34]. The model was populated with population estimates from the United Nations 2019 revision, which provide the number of girls alive in each year and age over the lifetimes of the birth cohorts included in the analysis [44]. For each single year of age and calendar year, the number of girls alive was multiplied by age-specific cervical cancer incidence and mortality rates to project the expected number of cervical cancer cases, deaths, and disability-adjusted life years (DALYs), both with and without vaccination. The model also estimated the costs of vaccination and the healthcare costs related to cervical cancer treatment in each scenario.

The primary outcome measure was the cost (USD) per DALY averted from a government perspective, accounting for all costs and benefits aggregated over the seventeen cohorts of vaccinated girls (2019–2035). DALYs were chosen because they combine years of life lost due to premature death and years lived with disability, enabling consistent comparison of health effects across diseases. All future costs and health outcomes were discounted at 3% per year to account for the time value of money and to reflect the preference for immediate benefits as well as the opportunity cost of investing current capital. This discount rate is commonly applied in economic evaluations of immunization programs, including HPV vaccination [32, 28, 14].

Senegal does not have a country-specific willingness-to-pay (WTP) threshold for determining whether a health intervention is cost-effective. Previous studies recommended that countries with a low Human Development Index adopt a threshold below 100% of GDP per capita, based on revealed WTP estimates from several low and middle-income countries [33, 35]. Another study from Kenya estimated a WTP threshold ranging between 0% and 50% of national GDP per capita [32]. Given the uncertainty surrounding the appropriate threshold for Senegal, the cost per disability-adjusted life year (DALY) averted was expressed as a percentage of the national GDP per capita (in USD) to facilitate interpretation of results. Therefore, we estimated the probability that the HPV vaccine would be cost-effective across a range of alternative WTP thresholds, from 0 to 0.3 times the national GDP per capita equivalent to up to USD 450, based on a GDP per capita of USD 1501 in 2019 [1].

### 2.3 Disease burden

Input data for disease burden are summarised in Supplementary Table 4. We used age-specific cervical cancer incidence and mortality rates estimated for Senegal by GLOBOCAN for the year 2022 [24], assuming these rates remain constant over time in the absence of vaccination or changes to current screening practices. Cancer staging definitions followed the International Federation of Gynecology and Obstetrics (FIGO) system [6]. Based on previously reported estimates for Senegal, we assumed the distribution of cases by cancer stage to be 33.3% local, 44.2% regional, and 22.5% distant [26]. Disability weights were drawn from the Global Burden of Disease project and reflect the time lived with cervical cancer at each stage [38]. Stage-specific five-year survival rates were derived from data reported in a recent study from Kenya [32], resulting in estimated average five-year survival rates in Senegal of 50.3%, 20.5%, and 0.0% for local, regional, and distant cervical cancer, respectively. For all model parameters without reported uncertainty intervals, we applied a ± 20% variation around the central estimate to define plausible ranges for use in the uncertainty analysis [20].

### 2.4 Healthcare costs

We assumed that all women represented in the GLOBOCAN incidence estimates [24] would be diagnosed and receive treatment. Therefore, the average cost of cervical cancer treatment was applied uniformly. Our cost calculations included only the direct costs of cervical cancer treatment, excluding expenses related to the screening and management of precancerous lesions. Indirect costs, such as lost wages for patients and their families, as well as broader government-borne expenditures—including health system costs related to personnel, hospital logistics, and infrastructure were also excluded.

Direct treatment-related costs were derived from a cost analysis conducted at the Joliot Curie Institute of Aristide Le Dantec Hospital in Dakar [11], which estimated medical costs per procedure for patients diagnosed with cervical cancer in Senegal. Treatment costs vary based on cancer stage, chemotherapy and radiotherapy protocols, and the number of diagnostic and follow-up laboratory and imaging tests required. The direct medical cost of cervical cancer treatment was estimated to range from USD 1,495.15 to USD 10,662.97, with an average of USD 3,713.45.

The total healthcare cost at a given stage of cervical cancer was calculated as the sum of all diagnostic and treatment procedures undertaken. Stage-specific treatment costs included expenditures for diagnosis, chemotherapy, radiotherapy, surgery (simple or radical hysterectomy), medications, medical consumables, inpatient care, and radiological investigations. On average, patients treated with chemotherapy alone incurred a cost of USD 1,187.76, those undergoing surgery USD 963.77, and those receiving radiotherapy USD 272.79 [11]. For local stage, costs included initial examination, surgery, and radiotherapy. Stage III disease required diagnosis, radiotherapy, chemotherapy, and medications. Distant-stage treatment involved radiotherapy and chemotherapy, followed by inpatient care and potential surgical intervention. Based on these components, we estimated the total cost of cervical cancer treatment to be USD 1,517.35 for local stage, USD 2,526.23 for regional stage, and USD 4,114.16 for distant stage, with an assumed average treatment cost of USD 3,713.45 across all stages. Inputs for healthcare costs are detailed in Supplementary Table 5.

### 2.5 Vaccine Coverage and Efficacy

Vaccine coverage estimates are presented in the Introduction for the period 2019–2024 and in Table 1 for 2025– 2035. In 2019, first- and second-dose coverage rates were estimated at 89% and 25%, respectively. However, coverage declined in 2020 to 47% for the first dose and 32% for the second dose. For the period 2022–2024, we assumed coverage levels of 47% for the first dose and 25% for the second dose, based on WHO immunization data [49]. For the period 2025–2030, we assumed a gradual scale-up, starting at 60% in 2025 and reaching 90% by 2030 for the first dose. These assumptions are informed by a school-based HPV vaccine pilot demonstration project conducted in Senegal from 2014 to 2016 [39], and are consistent with the WHO global target of achieving 90% coverage among girls.

HPV type distribution was based on a cross-sectional cervical cancer surveillance study conducted in Senegal between 2013 and 2017 [26]. The most common HPV types identified were 16 (100%), 18 (83%), and 45 (33%). Other prevalent types included 33 (31%), 59 (28%), 35 (12%), and 31 (11%). As the total proportion exceeded 100% due to co-infections, we rescaled the values to fit within a 100% envelope for analysis (Figure 2b). After rescaling, the most prevalent types were HPV 16 (31.75%), 18 (26.35%), 45 (10.48%), and 33 (9.84%).

**Figure 1:**
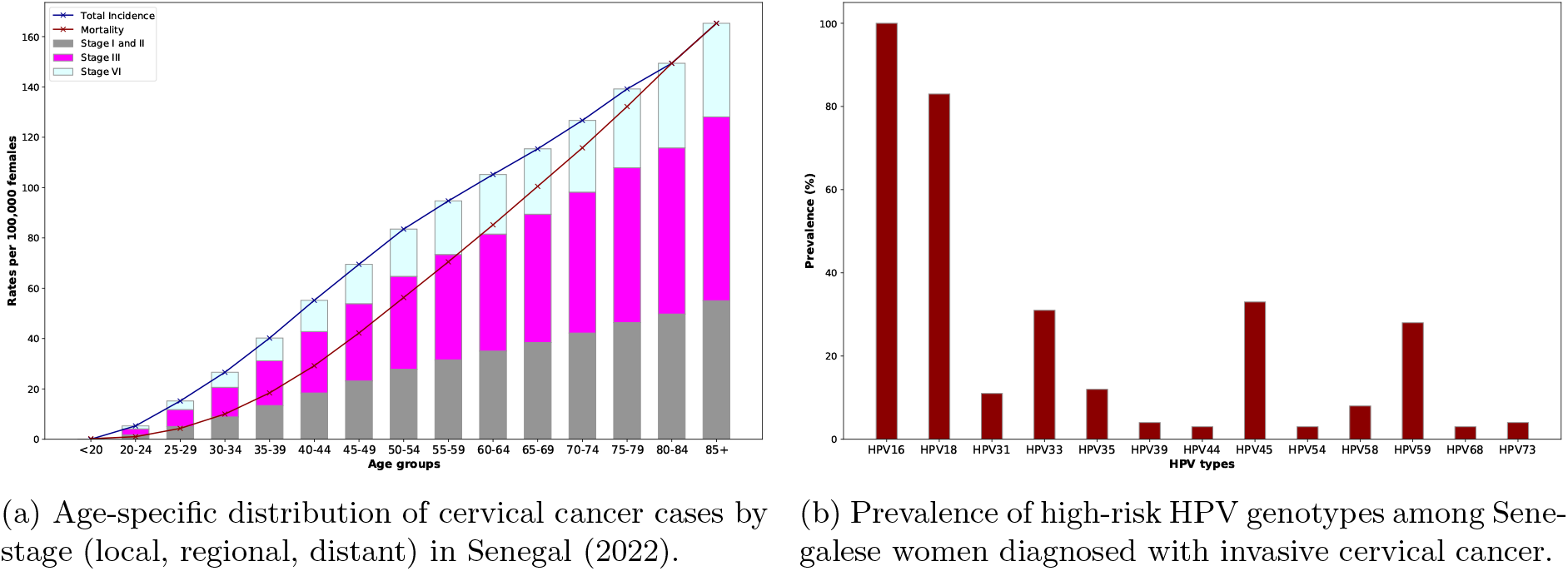
Burden of cervical cancer in Senegal: (a) stage-specific incidence by age group, and (b) distribution of high-risk HPV genotypes in invasive cervical cancer cases.

**Figure 2:**
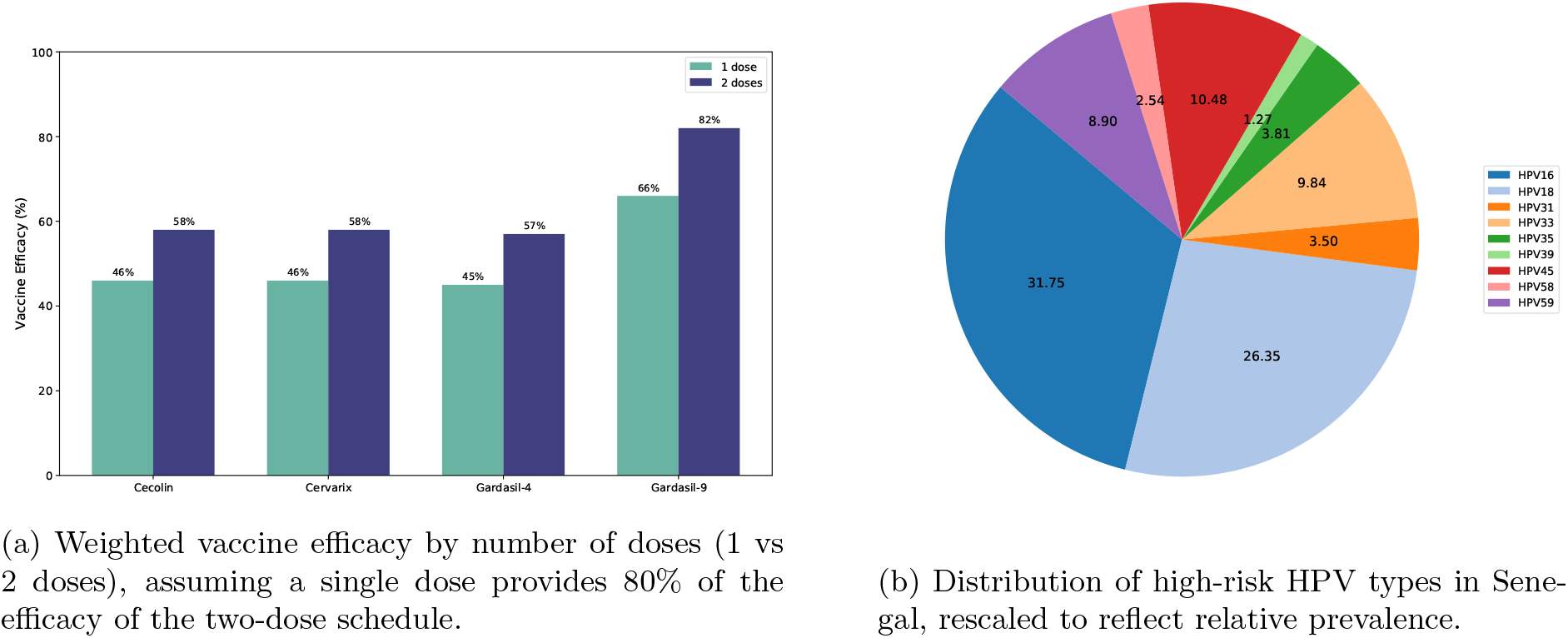
HPV vaccine efficacy by product and high-risk HPV type distribution in Senegal.

Vaccine-type efficacy estimates for the full (two-dose) vaccination schedule were obtained from Qiao et al. [37] for Cecolin, Apter et al. [2] for Cervarix, and Ault et al. [4] and Garland et al. [17] for Gardasil-4. Additional efficacy data for Gardasil-9 were taken from Huh et al. [22]. Weighted efficacy values for each vaccine product were calculated by multiplying the type-specific efficacy by the proportion of cervical cancer cases attributable to each HPV type in Senegal (Figure 2a). In the base-case analysis, we assumed that a single dose of the HPV vaccine would provide 80% (range: 70%–100%) of the total efficacy associated with two doses [37].

### 2.6 Vaccination Program Costs

Input data for vaccine program costs are summarized in Supplementary Table 6. A major cost driver for HPV vaccination programs in Africa is the vaccine price itself. However, many Gavi-supported countries benefit from negotiated reduced prices. Currently, Gavi-negotiated prices are USD 2.90 for Cecolin, USD 5.18 for Cervarix, and USD 4.50 for Gardasil-4 [19, 50]. Without Gavi support, vaccine costs represent a substantial financial barrier to HPV vaccination programs across the continent. Consequently, countries such as Senegal must develop sustainable long-term funding strategies to maintain vaccine access when Gavi support eventually phases out.

By 2030, the government is assumed to fully finance HPV vaccines as Gavi support phases out. Gardasil-9 is not yet available through Gavi, and the cost of self-procurement remains uncertain. Based on the lowest negotiated price for a non-Gavi country, we assume a cost of USD 25 per dose throughout the entire period (2019–2035) [50, 32]. To evaluate the financial impact, we also analyze a scenario without Gavi support, in which the government covers the full manufacturer’s vaccine price during the six-year period from 2030 to 2035.

In both scenarios with and without Gavi support, the government is expected to cover all costs related to vaccine wastage, international handling, international delivery, and other incremental health system expenses required to implement the vaccination program [41]. We assumed a 3% handling fee for all vaccines, based on the UNICEF fee applied to new and underused vaccines in least-developed countries [43], and a 10% international delivery fee to cover insurance and freight costs. Vaccine wastage was estimated at 5% for vaccines supplied in single-dose vials (Cecolin, Gardasil-4, and Gardasil-9) and 10% for Cervarix, which is supplied in two-dose vials. Unit costs per dose for syringes and safety boxes were USD 0.041 and USD 0.005, respectively, based on a retrospective cost evaluation conducted in Senegal [7].

The incremental health system costs include service delivery, planning, training, social mobilization, supervision and monitoring, cold chain (1%), and other minor expenses (*<*1%). Senegal leveraged existing cold chain and waste management infrastructure, so no additional investments were required when HPV vaccination was introduced nationally [9]. Service delivery represented the largest share of total weighted financial costs at 30%, followed by training at 42%. Planning accounted for 4%, social mobilization 14%, and supervision and monitoring 9%. Consequently, the incremental health system cost per dose was estimated at USD 3.07 for Gardasil-4 during the first year of introduction (2019) [7]. Assuming the same cost structure for other vaccines, with only vaccine price varying, the incremental health system cost remains USD 3.07 per dose. The recurrent cost per dose from 2020 to 2028 was assumed to be USD 0.92, reflecting the 30% service delivery component.

### 2.7 Cost-effectiveness and sensitivity analysis

We analyzed the incremental costs and incremental disability-adjusted life years (DALYs) for each vaccination strategy compared to both the baseline scenario and the next best (i.e., lower-cost, non-dominated) strategy. The incremental cost-effectiveness ratio (ICER) was calculated as the ratio of the difference in costs to the difference in effectiveness (DALYs averted). Formally, the ICER represents the additional cost required to avert one additional DALY and is defined as:

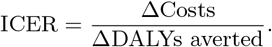

By plotting the cost-effectiveness frontier, which connects all non-dominated strategies, we identified the set of potentially optimal interventions. Following a recent global study proposing a new framework for estimating willingness-to-pay (WTP) thresholds for 174 countries [35], we adopted a WTP threshold of 30% of Senegal’s GDP per capita in 2019, equivalent to USD 450.

Probabilistic sensitivity analyses were conducted to capture the combined uncertainty across all model parameters using 1000 Monte Carlo simulations. Furthermore, we evaluated how varying the price of Gardasil-9 affects the ICER for two vaccination strategies: the least cost-effective option and the most costly option.

## 3 Results

If HPV vaccination coverage in Senegal remains at current levels with Gardasil-4 and no further improvements are made, it is projected that 130,204 girls will develop cervical cancer over their lifetimes, and 101,534 will eventually die from the disease (Table 2). Under this scenario, the total cost of cervical cancer-related treatment is estimated at USD 61 million over the lifetimes of 16 cohorts of 9-year-old girls, from the perspective of the government.

**Table 2:** Discounted health and economic outcomes of HPV vaccination strategies and baseline scenario over 17 birth cohorts from a government perspective (2019–2035).

| Outcomes | Health impact |  |  | Economic outcomes |  |  |
| --- | --- | --- | --- | --- | --- | --- |
|  | CC cases | CC deaths | DALYs | Vaccination costs | Treatment costs | Total costs |
| **Baseline** | **130,204** | **101,534** | **300,691** | **21,704,432** | **61,076,890** | **82,781,321** |
| Strategy 1 | 129,315 | 100,839 | 298,609 | 11,365,012 | 60,653,488 | 72,018,500 |
| Strategy 2 | 129,315 | 100,839 | 252,730 | 21,620,548 | 60,653,488 | 82,274,036 |
| Strategy 3 | 109,714 | 85,519 | 252,730 | 73,077,845 | 51,323,868 | 124,401,713 |
| Strategy 4 | 133,514 | 104,083 | 307,919 | 16,357,370 | 62,536,921 | 78,894,291 |
| Strategy 5 | 132,625 | 103,389 | 305,837 | 9,175,469 | 62,113,520 | 71,288,989 |
| Strategy 6 | 132,625 | 103,389 | 305,837 | 16,861,899 | 62,113,520 | 78,975,419 |
| Strategy 7 | 114,127 | 88,919 | 262,367 | 59,225,034 | 53,270,577 | 112,495,611 |
| Strategy 8 | 120,863 | 94,367 | 280,570 | 27,244,736 | 57,013,766 | 84,258,502 |
| Strategy 9 | 119,787 | 93,529 | 278,085 | 13,516,734 | 56,508,978 | 70,025,712 |
| Strategy 10 | 119,787 | 93,529 | 278,085 | 26,433,123 | 56,508,978 | 82,942,101 |
| Strategy 11 | 96,137 | 75,103 | 223,483 | 86,517,808 | 45,418,049 | 131,935,857 |
| Strategy 12 | 125,097 | 97,625 | 289,783 | 20,213,841 | 58,874,516 | 79,088,357 |
| Strategy 13 | 124,021 | 96,787 | 287,298 | 10,705,711 | 58,369,728 | 69,075,439 |
| Strategy 14 | 124,021 | 96,787 | 287,298 | 20,244,602 | 58,369,728 | 78,614,330 |
| Strategy 15 | 101,782 | 79,446 | 235,767 | 68,833,968 | 47,899,050 | 116,733,018 |
| Strategy 16 | 109,610 | 85,698 | 255,994 | 33,368,484 | 52,049,658 | 85,418,142 |
| Strategy 17 | 108,313 | 84,690 | 253,027 | 16,164,731 | 51,447,534 | 67,612,265 |
| Strategy 18 | 108,313 | 84,690 | 253,027 | 32,024,721 | 51,447,534 | 83,472,255 |
| Strategy 19 | 79,808 | 62,524 | 187,825 | 103,479,544 | 38,215,226 | 141,694,770 |
| Strategy 20 | 115,167 | 89,976 | 268,098 | 24,198,764 | 54,494,421 | 78,693,185 |
| Strategy 21 | 113,871 | 88,968 | 265,131 | 12,477,891 | 53,892,297 | 66,370,188 |
| Strategy 22 | 113,871 | 88,968 | 265,131 | 23,932,740 | 53,892,297 | 77,825,037 |
| Strategy 23 | 87,218 | 68,228 | 203,963 | 80,254,579 | 41,474,910 | 121,729,489 |

A comprehensive analysis of 23 HPV vaccination strategies in Senegal over the period 2019–2035 highlights the potential health and economic impacts of different vaccine products, dose schedules, and coverage levels ( Table 2 and Table 3). In the evaluated scenarios, vaccine procurement benefits from GAVI support from 2019 to 2030, while from 2030 onwards, the government transitions to purchasing vaccines at manufacturer prices. Under these conditions, most alternative strategies substantially outperform the current national approach with two doses of Gardasil 4 at constant coverage in reducing the burden of cervical cancer. Bivalent vaccines such as Cecolin and Cervarix, administered as either one or two doses, provide substantial health benefits and cost savings, particularly in scenarios with increased coverage. For example, Strategies 17 and 21, which involve Cecolin under the WHO 90% coverage target, lead to reductions in disability-adjusted life years (DALYs) of 15.85% and 11.83%, respectively, while also being cost-saving. Similar trends are observed with Cervarix in Strategies 18 and 22. One-dose schedules are generally more cost-effective, achieving strong health outcomes while reducing programmatic costs. In comparison, strategies based on GARDASIL 9, such as Strategies 3, 7, 11, 15, 19, and 23, result in the highest absolute DALY reductions, reaching up to 37.5%, but are associated with considerably higher costs and less favorable incremental cost-effectiveness ratios (ICERs). These findings indicate that bivalent vaccines administered through simplified one-dose schedules with broader coverage offer the most economically favorable strategy for HPV prevention in Senegal.

**Table 3:** Incremental and cost-effectiveness outcomes by strategy.

| Strategy | Incremental effectiveness (DALYs) | Incremental Costs (USD) | DALYs Reduction (%) | Net Cost Ratio | ICER (USD/DALY) |
| --- | --- | --- | --- | --- | --- |
| Baseline | – | – | – | – | – |
| Strategy 1 | 2,082 | -10,762,821 | 0.69 | -0.13 | Cost saving |
| Strategy 2 | 47,961 | -507,285 | 15.95 | -0.01 | Cost saving |
| Strategy 3 | 47,961 | 41,620,392 | 15.95 | 0.50 | 868 |
| Strategy 4 | -7,228 | -3,887,030 | -2.40 | -0.05 | 538 |
| Strategy 5 | -5,146 | -11,492,332 | -1.71 | -0.14 | 2,233 |
| Strategy 6 | -5,146 | -3,805,902 | -1.71 | -0.05 | 740 |
| Strategy 7 | 38,324 | 29,714,290 | 12.75 | 0.36 | 775 |
| Strategy 8 | 20,121 | 1,477,181 | 6.69 | 0.02 | 73 |
| Strategy 9 | 22,606 | -12,755,609 | 7.52 | -0.15 | Cost saving |
| Strategy 10 | 22,606 | 160,780 | 7.52 | 0.00 | 7 |
| Strategy 11 | 77,208 | 49,154,536 | 25.68 | 0.59 | 637 |
| Strategy 12 | 10,908 | -3,692,964 | 3.63 | -0.04 | Cost saving |
| Strategy 13 | 13,393 | -13,705,882 | 4.45 | -0.17 | Cost saving |
| Strategy 14 | 13,393 | -4,166,991 | 4.45 | -0.05 | Cost saving |
| Strategy 15 | 64,924 | 33,951,697 | 21.59 | 0.41 | 523 |
| Strategy 16 | 44,697 | 2,636,821 | 14.87 | 0.03 | 59 |
| Strategy 17 | 47,664 | -15,169,056 | 15.85 | -0.18 | Cost saving |
| Strategy 18 | 47,664 | 690,934 | 15.85 | 0.01 | 14 |
| Strategy 19 | 112,866 | 58,913,449 | 37.53 | 0.71 | 522 |
| Strategy 20 | 32,593 | -4,088,136 | 10.84 | -0.05 | Cost saving |
| Strategy 21 | 35,560 | -16,411,133 | 11.83 | -0.20 | Cost saving |
| Strategy 22 | 35,560 | -4,956,284 | 11.83 | -0.06 | Cost saving |
| Strategy 23 | 96,728 | 38,948,168 | 32.16 | 0.47 | 403 |

**Table 4:** Input parameters for estimating cervical cancer disease burden in Senegal.

| Parameter | Value | Source |
| --- | --- | --- |
| **Annual rate of cervical cancer cases per 100,000 females** |  |  |
| < 20 years | 0.0 | GLOBOCAN 2022 [24] |
| 20–24 years | 5.3 | - |
| 25–29 years | 15.2 | - |
| 30–34 years | 26.6 | - |
| 35–39 years | 40.2 | - |
| 40–44 years | 55.2 | - |
| 45–49 years | 69.5 | - |
| 50–54 years | 83.5 | - |
| 55–59 years | 94.7 | - |
| 60–64 years | 105.2 | - |
| 65–69 years | 115.4 | - |
| 70–74 years | 126.7 | - |
| 75–79 years | 139.2 | - |
| 80–84 years | 149.4 | - |
| 85+ years | 165.3 | - |
| **Annual rate of cervical cancer deaths per 100,000 females** |  |  |
| < 20 years | 0.0 | GLOBOCAN 2022 |
| 20–24 years | 0.21 | - |
| 25–29 years | 1.0 | - |
| 30–34 years | 4.3 | - |
| 35–39 years | 10.0 | - |
| 40–44 years | 18.4 | - |
| 45–49 years | 29.2 | - |
| 50–54 years | 42.2 | - |
| 55–59 years | 56.3 | - |
| 60–64 years | 70.5 | - |
| 65–69 years | 100.5 | - |
| 70–74 years | 115.8 | - |
| 75–79 years | 132.2 | - |
| 80–84 years | 149.4 | - |
| 85+ years | 165.3 | - |
| **% distribution of cervical cancer by severity** |  |  |
| stage I and II | 33.3 % | [26] |
| stage III | 44.2 % | - |
| stage IV | 22.5 % | - |
| **% High-risk HPV genotypes distribution among invasive cervical cancer cases** |  |  |
| 16/18 | 58.1 % | [26] |
| 16/18/31/33/45/52/58 | 84.46 % | - |
| **Disability weights for cervical cancer by stage** |  |  |
| Local | 0.288 | Diagnosis and primary therapy phase of cervical cancer [38] |
| Regional | 0.45 | Metastatic phase - |
| Distant | 0.54 | Terminal phase - |
| **% cancer alive after 5 years** |  |  |
| Local | 50.3 % | [32] |
| Regional | 20.5 % | - |
| Distant | 0 % | - |

**Table 5:** Input parameters for health service costs from the government perspective.

| Parameter | Value | Source |
| --- | --- | --- |
| **CC treatment cost per case (government perspective)** |  |  |
| Local | 1517.35 | [11] |
| Regional | 2526.23 | - |
| Distant | 4114.16 | - |
| Average cancer treatment cost per case | 3713.45 | - |

**Table 6:** Input parameters for estimating HPV vaccine program costs.

| Parameter | Value | Source |
| --- | --- | --- |
| **Vaccine price, per dose (USD)** |  |  |
| Cecolin | 2.90 | [19] |
| Cervarix | 5.18 | - |
| Gardasil-4 | 4.5 | - |
| Gardasil-9 | 25 | [50, 32] |
| **Vaccine supplies, per dose (USD)** |  |  |
| Syringe price | 0.041 | [7] |
| Safety box price | 0,005 | - |
| International handling fee ( % of price) | 3 % | [43] |
| International delivery ( % of price) | 10 % | [7] |
| Wastage percentage | 5 % | - |
| **Incremental health system per year, per dose (USD)** |  |  |
| 2019 | 3.07 | [7] |
| 2020-2035 | 0.92 | - |

### 3.1 Cost-effectiveness of all HPV vaccination strategies

The incremental disability-adjusted life years (DALYs) and costs associated with 23 HPV vaccination strategies, compared to the baseline, are presented in Table 3 and Figure 3. These strategies show a wide range of impacts, with incremental costs ranging from a reduction of USD 16.41 million to an increase of USD 58.91 million, and health outcomes varying from a loss of 7,228 DALYs to a gain of 112,866 DALYs. Among them, 15 strategies were cost-saving, simultaneously improving health outcomes and reducing healthcare expenditures. Strategies involving Cecolin and Gardasil 9 were consistently among the most effective and non-dominated. Strategy 19, using Gardasil 9 with 90% coverage, achieved the greatest health benefit, averting 112,866 DALYs, though at a higher cost of USD 142 million and with an incremental cost-effectiveness ratio (ICER) of USD 522 per DALY averted. In contrast, Strategy 17, which uses Cecolin with 90% coverage, averted 47,664 DALYs while saving more than USD 15 million.

**Figure 3:**
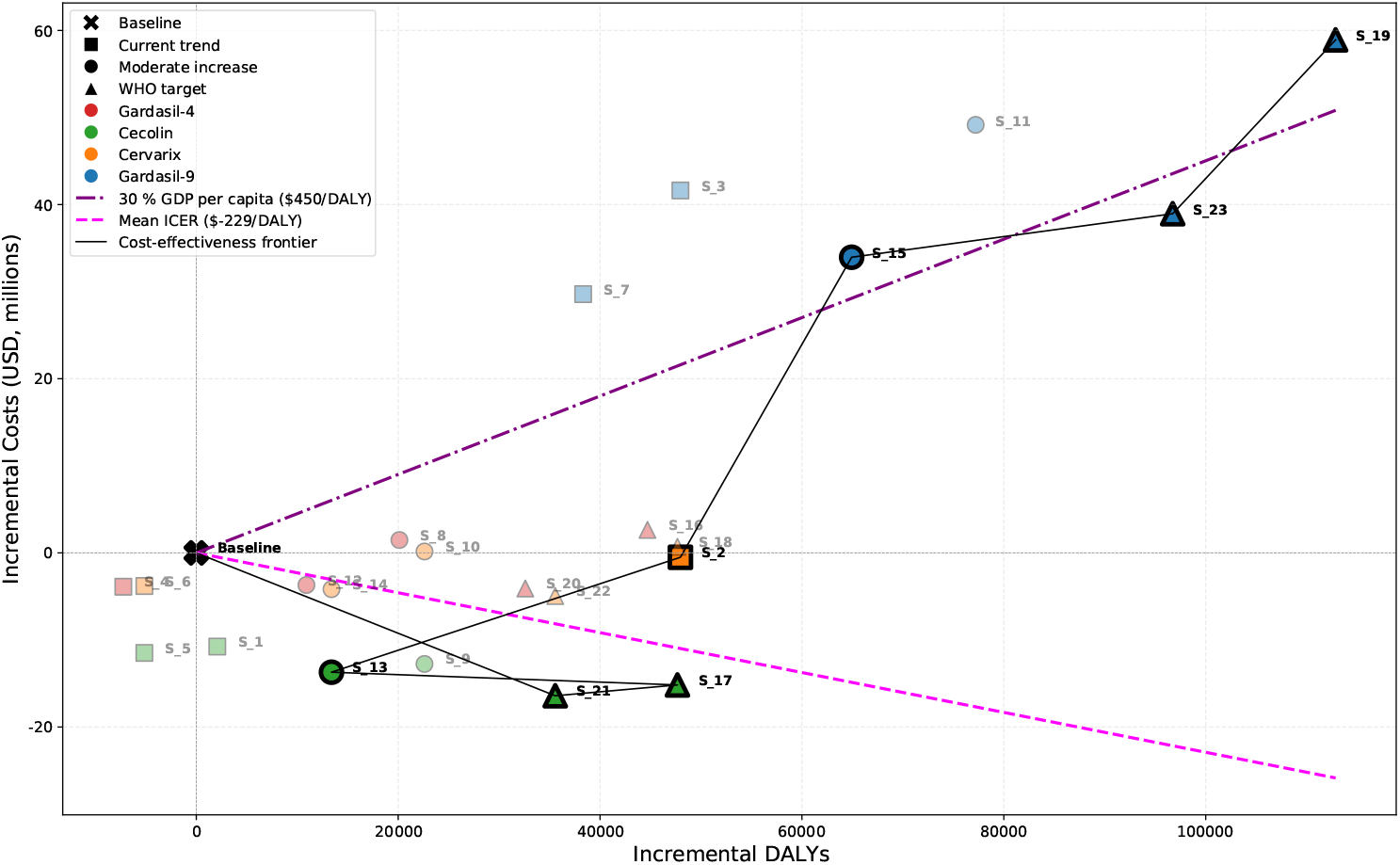
Cost-effectiveness frontier for all HPV vaccination strategies (100,000 cohort members). (A) Curent trend. (B) Moderate increase. (C) WHO target .Names of the strategies located on the cost-effectiveness frontier compared with the lower-cost non-dominated strategy are shown.

Strategies based on single-dose schedules using affordable vaccines, such as Cecolin or Cervarix, and implemented with expanded coverage, consistently delivered strong health gains and high economic efficiency. Some strategies with very low ICERs, such as Strategy 10 (USD 7 per DALY) and Strategy 18 (USD 14 per DALY), represent highly efficient public health investments. Conversely, a small number of strategies including Strategies 4, 5, and 6 resulted in negative health outcomes, suggesting that reducing the number of doses without increasing coverage may diminish program effectiveness. These findings underscore the importance of both expanding coverage and selecting cost-effective vaccine products to maximize the health and economic benefits of HPV vaccination programs in Senegal.

### 3.2 Optimal vaccination strategies

Most HPV vaccination strategies in Senegal are projected to prevent a substantial number of cervical cancer cases and related deaths compared to the baseline scenario. However, a few strategies result in worse health outcomes, including a higher number of cases and deaths, primarily due to insufficient coverage or reduced dosing without compensatory gains. These strategies are considered dominated, as they are both less effective and more costly than at least one alternative. In contrast, non-dominated strategies define the efficiency frontier, representing the set of interventions that offer the best health outcomes for their cost. Strategies located on this frontier are considered reasonably efficient and provide valuable guidance for policy prioritization (Figure 3).

Seven HPV vaccination strategies were identified along the cost-effectiveness frontier, most of which involved Cecolin, administered as either a one or two-dose schedule across all evaluated coverage levels. These strategies consistently emerged as the most cost-saving options. One-dose Gardasil-9 strategies under moderate and high (WHO target) coverage levels also appeared efficient, with ICERs of USD 521 and USD 403 per DALY averted, respectively. Applying the threshold of 30% of Senegal’s GDP per capita (USD 450), only five strategies met cost-effectiveness criteria: all Cecolin-based options and the one-dose Gardasil-9 strategy under 90% coverage. Cecolin interventions were projected to reduce cervical cancer incidence by 12% to 16% over the cohort’s lifetime compared to the baseline. In contrast, the one-dose Gardasil-9 strategy at WHO coverage could reduce incidence by up to 32%, while being associated with a 47 % increase in costs. Although this strategy lies just below the cost-effectiveness threshold and remains on the frontier, the two-dose Gardasil-9 approach under the same coverage level exceeds the threshold (ICER: USD 522 per DALY averted) and is therefore not considered cost-effective in this setting (Table 3, Figure 4).

**Figure 4:**
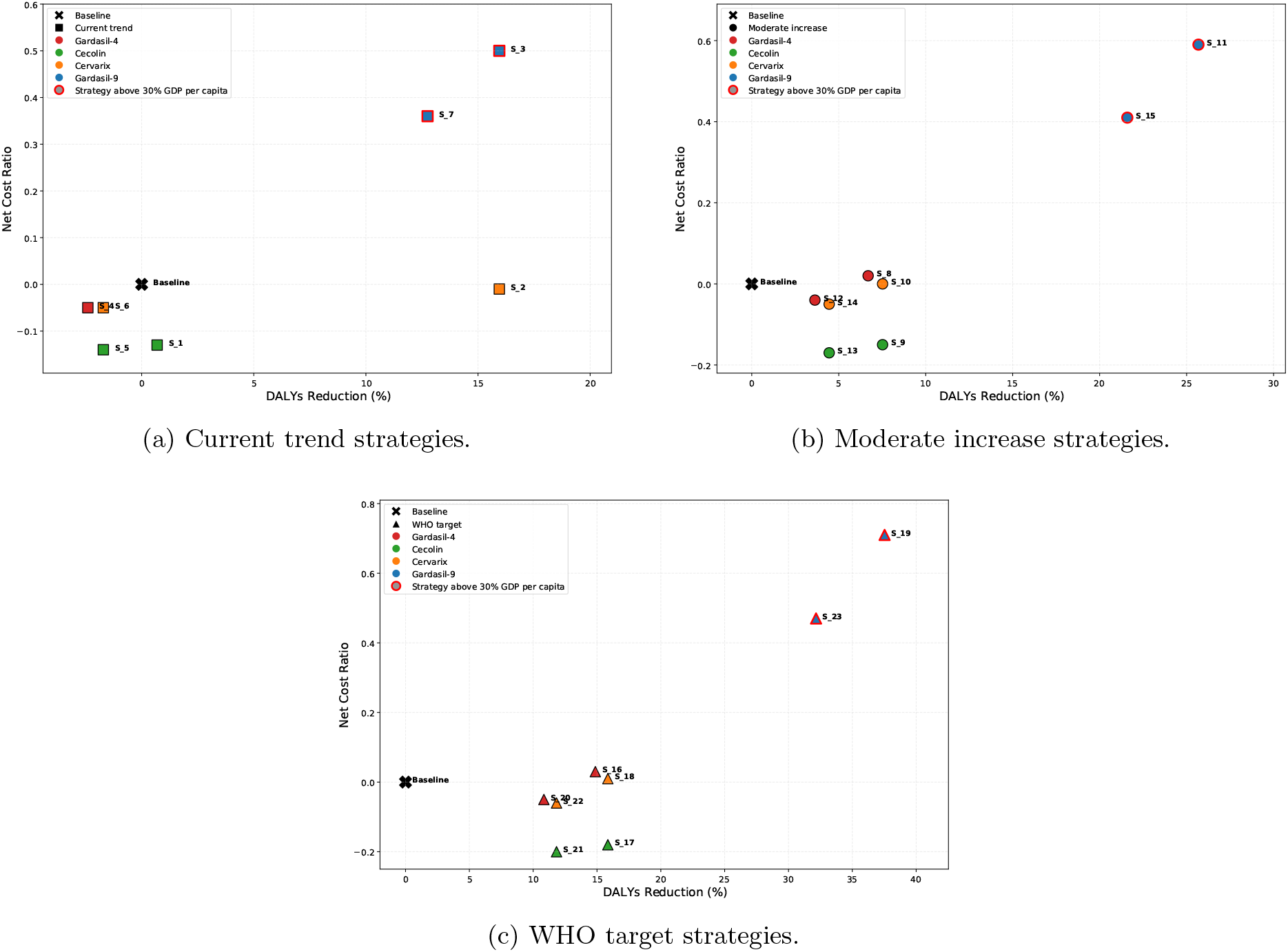
Cost-effectiveness plane: change in costs and number of DALYs per woman obtained when comparing a new vaccination strategy to the baseline.

These findings highlight Cecolin as a highly efficient and cost-saving option, while indicating that Gardasil-9 could generate greater health gains, albeit with substantially higher investment requirements. This contrast is illustrated in Figure 5, where Strategy 21 (Cecolin, one dose, WHO coverage) achieves substantial case reductions compared to the baseline, with annual undiscounted program costs exceeding USD 2 million at 90 % coverage, whereas Strategy 23 (Gardasil-9, one dose, WHO coverage) is identified as the most cost-effective among non-dominated strategies but would surpass USD 7 million annually once Senegal transitions from Gavi support.

**Figure 5:**
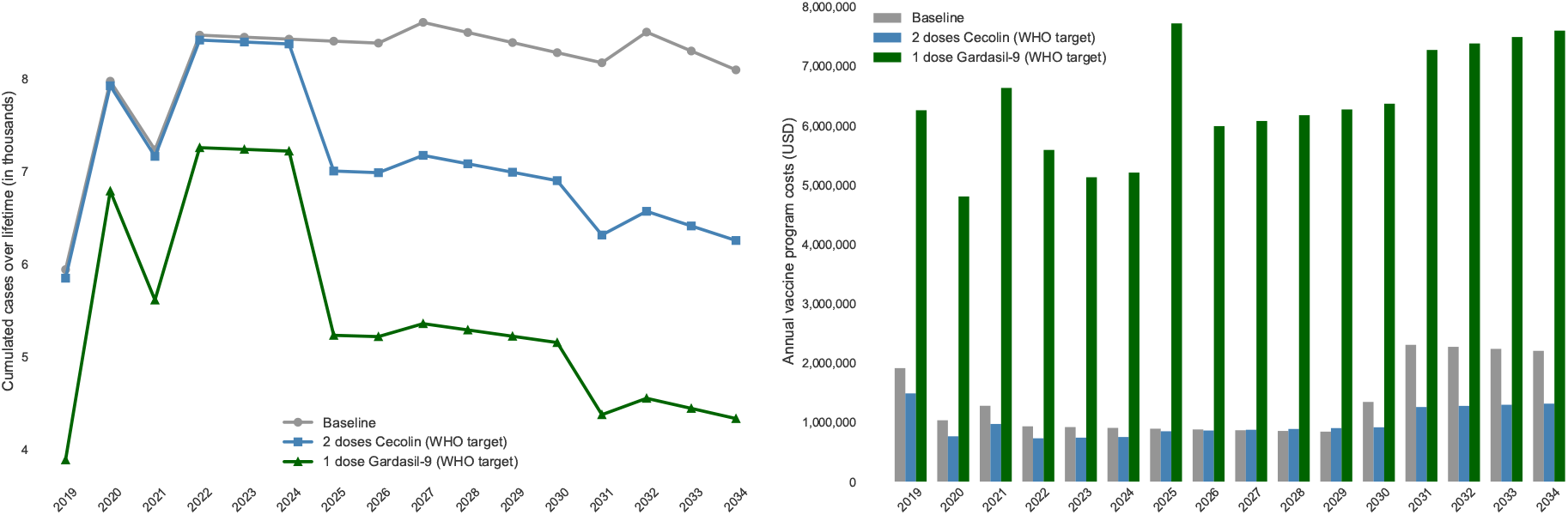
Cumulated cases over lifetime and undiscounted vaccine program costs (including incremental health system costs) by calendar year and type of HPV vaccine product (most saving and less most cost effective strategies).

### 3.3 Impact of lowering the Gardasil-9 price on the optimal vaccination strategy

Given the substantial health benefits associated with Gardasil-9, we explored how reductions in its price could influence the cost-effectiveness of one and two-dose vaccination strategies under the WHO 90% coverage scenario. As shown in Figure 6, the one-dose Gardasil-9 strategy (Strategy 23) is currently cost-effective, with an ICER of USD 403 per DALY averted, which is just below the willingness-to-pay threshold (USD 450 per DALY averted). Lowering the vaccine price further improves its cost-effectiveness, making it an increasingly favorable option. In contrast, the two-dose Gardasil-9 strategy (Strategy 19) exceeds the threshold at current prices, with an ICER of USD 522 per DALY averted, and is therefore not considered cost-effective in the present context. However, this strategy becomes cost-effective if the price per dose falls below USD 20, illustrating the critical role of vaccine pricing in determining economic feasibility. Overall, the viability of Gardasil-9, particularly in two-dose schedules, is highly sensitive to pricing, reinforcing the importance of securing affordable procurement to ensure sustainable and efficient HPV vaccination in Senegal.

**Figure 6:**
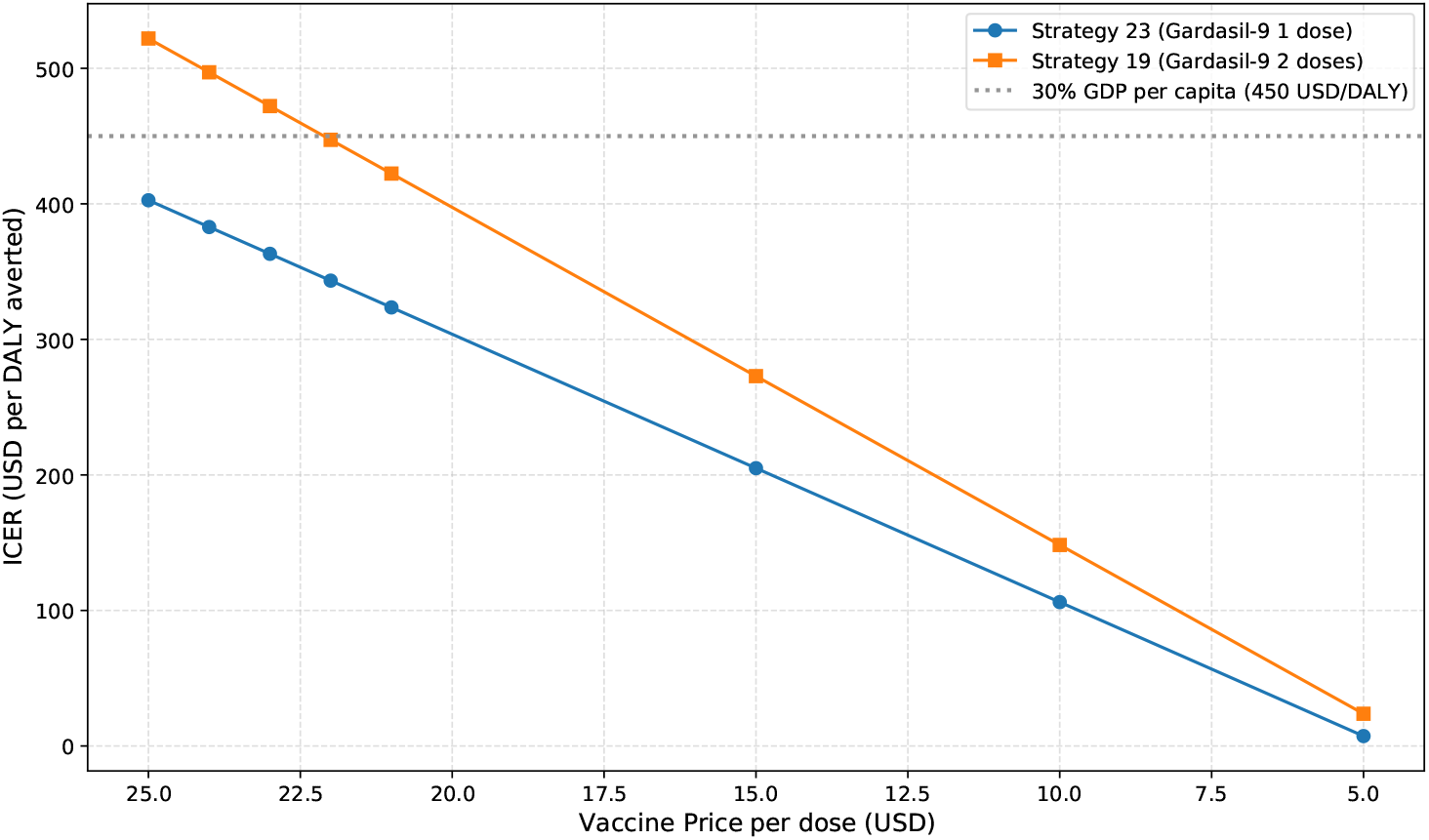
The effect of Gardasil-9 vaccine price per dose on ICER (USD / DALY averted) of the most efffective strategies (19 and 23).

### 3.4 Sensitivity analysis

To assess the robustness of our findings and account for parameter uncertainty, we conducted probabilistic sensitivity analyses (PSA) for two representative strategies: Strategy 21 (Cecolin, 1 dose, WHO coverage), identified as the most cost-saving option, and Strategy 23 (Gardasil-9, 1 dose, WHO coverage), identified as the most cost-effective among non-dominated strategies (Figure 5). For each, 500 Monte Carlo simulations were run, simultaneously varying all epidemiological, coverage, and cost parameters using PERT-Beta distributions. Vaccine prices were kept constant to isolate the influence of other variables. The outputs were used to generate cost-effectiveness acceptability curves (CEACs), which estimate the probability that a given strategy is cost-effective across a range of willingness-to-pay (WTP) thresholds (Figure 7).

**Figure 7:**
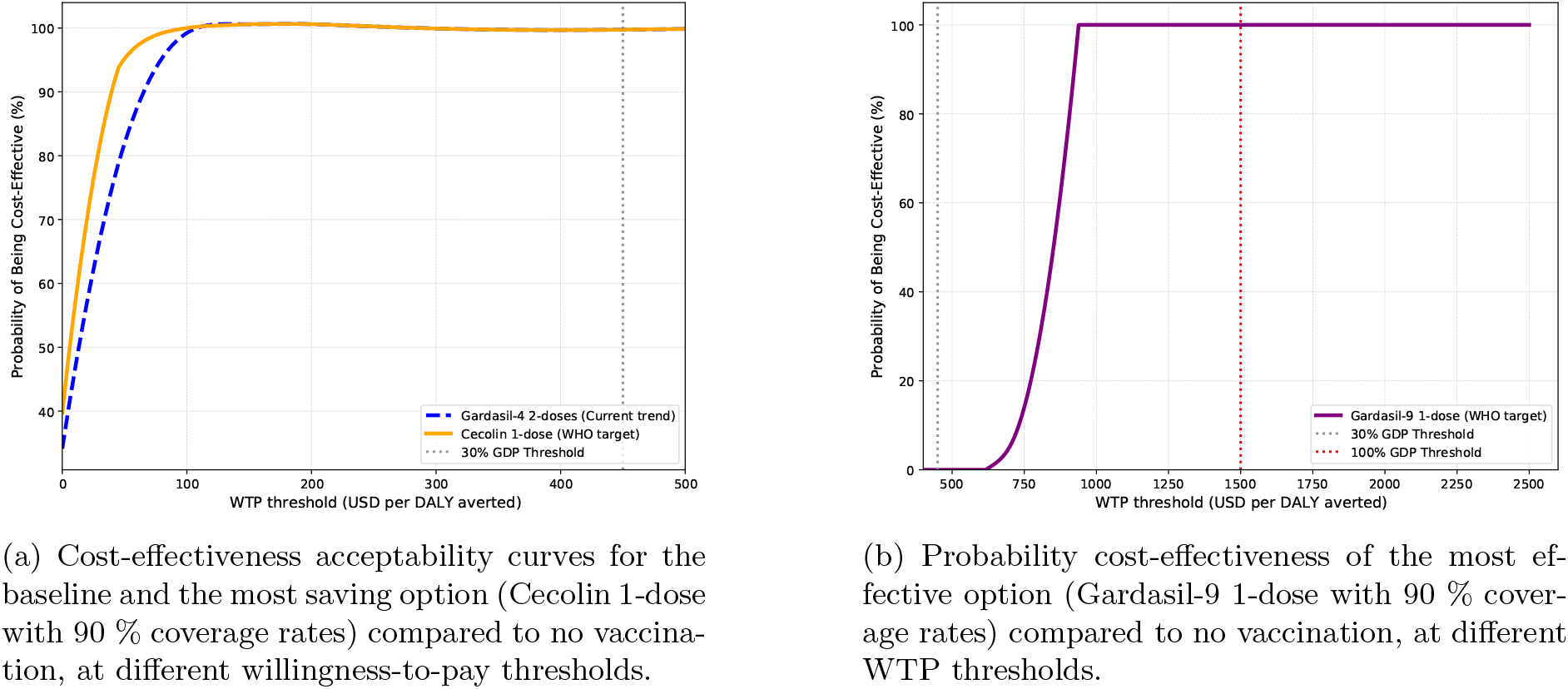
Cost-effectiveness acceptability curves for three vaccination strategies: Baseline, Cecolin and Gardasil-9 1-dose (WHO target) compared to no vaccination, at different WTP thresholds.

Cecolin demonstrates a highly robust cost-effectiveness profile, reaching a 100% probability of being cost-effective at a WTP threshold of approximately USD 100 per DALY averted (less than 7% of Senegal’s GDP per capita). Notably, while the acceptability curves for Cecolin and the baseline strategy (Gardasil-4, 2 doses) tend to converge at higher WTP thresholds, they begin to diverge sharply below USD 100, where Cecolin consistently demonstrates a higher probability of being cost-effective (Figure 7a). This suggests that, under more constrained budget scenarios, Cecolin offers a significantly more favorable economic profile compared to the baseline.

In contrast, Gardasil-9 achieves an 80% probability of being cost-effective only at a WTP threshold of USD 800 (approximately 53% of GDP per capita), and reaches 100% probability only at thresholds above USD 900 (Figure 7b). These results confirm that, while Gardasil-9 is associated with greater health benefits, its economic performance is more sensitive to available resources. Under constrained budget conditions, Cecolin remains the most reliable and efficient choice, while broader adoption of Gardasil-9 would require either increased financial capacity or lower vaccine prices.

## 4 Discussion

This study provides a comprehensive evaluation of the health and economic impacts of alternative HPV vaccination strategies in Senegal, examining different vaccine types, dosing regimens, and coverage levels. Using a model-based approach, we found that bivalent HPV vaccine strategies, particularly those employing simplified single-dose schedules with expanded coverage, were highly cost-effective relative to a threshold of USD 450 per DALY averted (based on GDP per capita), achieving a 16% reduction in cervical cancer risk compared to the current national two-dose Gardasil-4 program. In contrast, strategies involving Gardasil-9, while yielding the largest absolute health benefits (up to 112,866 DALYs averted), were associated with substantially higher programmatic costs and less favorable incremental cost-effectiveness ratios (ICERs). Five strategies (mainly those based on Cecolin or single-dose Gardasil-9) at WHO-recommended coverage levels, were considered efficient under the national willingness-to-pay threshold of USD 450 per DALY averted. These optimal strategies remained robust across multiple sensitivity and scenario analyses. At lower willingness-to-pay thresholds (e.g., 7% of GDP per capita or USD 100), the one-dose Cecolin strategy with 90% coverage (Strategy 21) emerged as the most cost-effective, though it offered more limited reductions in cervical cancer risk (16%). Gardasil-9 (Strategy 23) also proved cost-effective at current prices, with an ICER of USD 403 per DALY averted, slightly below the defined threshold. However, when the vaccine unit cost was reduced to USD 20 in the base case, the two-dose Gardasil-9 strategy at WHO coverage became one of the most optimal options, highlighting the pivotal role of procurement costs in determining the economic feasibility of higher-cost vaccines.

Determining whether an intervention should be included in a health benefits package requires establishing a benchmark for health opportunity costs, commonly known as a cost-effectiveness threshold. The WHO Commission on Macroeconomics and Health has proposed that interventions with incremental cost-effectiveness ratios (ICERs) below a country’s GDP per capita be considered “very cost-effective” and those below three times GDP per capita as “cost-effective”. However, recent research has challenged the appropriateness of this approach, suggesting that it may result in the displacement of more efficient interventions by those generating fewer health benefits. In the absence of a formal, empirically derived threshold for Senegal, we examined willingness-to-pay (WTP) thresholds ranging from 0 to 0.3 times the national GDP per capita, informed by WTP estimates observed in several low- and middle-income countries. Importantly, beyond considerations of economic efficiency, equity remains a critical factor in determining which services should be prioritized and made universally accessible.

Senegal was the first Gavi-supported country in West Africa to integrate the HPV vaccine into its routine immunization program. While Gavi-funded pilot demonstration projects have provided valuable insights into delivering the vaccine to populations that do not routinely access health services, significant gaps remain in understanding the processes involved in national introduction and scale-up. These include challenges related to effective programmatic decision-making, resource allocation, implementation strategies, and key determinants for achieving equitable access.

Several studies have examined the cost-effectiveness of HPV vaccination strategies in low- and middle-income African countries. Michaeli et al. [30] reported that introducing the nonavalent vaccine alongside the existing screening program for bivalent HPV vaccination in South Africa was cost-effective, with the nonavalent vaccine emerging as the preferred strategy in 90.2% of simulations. In Burkina Faso, Kiendrébéogo et al. [27] estimated that HPV vaccination could prevent 37-72% of cervical cancer cases and deaths, identifying Cecolin as the most cost-effective option. They also highlighted that adopting a single-dose schedule and/or using alternative Gavi-supported vaccines could further improve cost-effectiveness. Similarly, Wondimu et al. [47] found that the 9-valent vaccine would be cost-effective in Ethiopia at a threshold of one GDP per capita, with an ICER of USD 107 per DALY averted, whereas the 4-valent vaccine did not meet this threshold. When comparing our results with those of previous studies, it is clear that, despite variations in methodology and assumptions, there is general agreement on the effectiveness and economic value of HPV vaccination in reducing the cervical cancer burden in African LMICs.

Several limitations should be acknowledged. First, although nationally representative demographic and epidemiological data were used, certain key parameters such as long term disease progression rates and vaccine efficacy were derived from international sources, potentially introducing uncertainty. Nonetheless, extensive sensitivity analyses were performed to address this issue. Findings from other studies underscore the importance of calibrating model inputs to better reflect local conditions, including diagnostic practices and country specific cost data [8]. While static models are widely accepted for evaluating the cost effectiveness of HPV vaccination, it is important to recognize the limitations in the current literature. These include reliance on non local data, assumptions of high vaccine uptake, and the omission of critical factors such as vaccine hesitancy, herd immunity, and HIV status. For example, current guidelines recommend an additional vaccine dose for HIV positive adolescents, which has significant implications for cost estimates and must be incorporated into any model intended for accurate policymaking [16].

Despite certain limitations, the findings provide timely and actionable insights for policymakers evaluating strategic adjustments to HPV vaccination in Senegal, particularly given that three major coverage scenarios were assessed. The consistent dominance of Cecolin-based strategies highlights a clear and cost-effective pathway for enhancing cervical cancer prevention efforts. Additionally, strategic price negotiations for Gardasil-9 could expand viable options with substantial health benefits. Future research should examine the long-term budgetary implications, equity impacts, and implementation feasibility to support effective national scale-up. From a policy standpoint, these results carry important implications. First, they advocate for a shift toward bivalent vaccines such as Cecolin, especially in the context of ongoing Gavi support and the country’s anticipated transition from donor assistance. The strong performance of one-dose regimens also offers significant advantages in terms of programmatic simplicity and reduced delivery costs, making them particularly well-suited for low-resource settings. Second, while Gardasil-9 provides broader protection, its cost-effectiveness is highly contingent on achieving significant price reductions and maintaining high coverage levels. Without these conditions, Cecolin-based approaches represent a more pragmatic and scalable solution for national HPV vaccination programs.

This analysis shows that several HPV vaccination strategies could provide substantial health and economic benefits for Senegal compared to the current national strategy based on constant coverage with Gardasil-4. In particular, bivalent vaccines such as Cecolin and Cervarix, delivered as a single-dose and scaled up to 90% coverage (in line with the WHO target), emerge as the most cost-effective options, yielding significant reductions in DALYs and even net savings for the health system. While strategies using Gardasil-9 achieve the largest absolute reductions in cervical cancer burden, their high cost makes them less attractive from an economic efficiency standpoint.

Assuming a single dose strategy had more favourable cost-effectiveness, reducing the cost per DALY averted by more than half. If costs are prohibitive, then a single dose strategy could be considered to reduce the cost. A further enhancement to the programme would be to include HPV vaccination for boys, but evaluation of this would require the use of a more complicated model, and was therefore outside the scope of our current study.

## 5 Conclusion

This study provides evidence that multiple HPV vaccination strategies can yield substantial health and economic benefits in Senegal, particularly as the country transitions from Gavi support. Among the evaluated options, single-dose bivalent vaccine strategies, especially those involving Cecolin at high coverage levels, consistently emerged as the most cost-effective and programmatically feasible. While Gardasil-9 offers the greatest absolute health gains, its cost-effectiveness is highly sensitive to vaccine pricing. These findings underscore the importance of aligning vaccine selection with both economic efficiency and implementation capacity to ensure sustainable and equitable cervical cancer prevention efforts in Senegal.

## Data Availability

All data produced are available online at global datasets.

## 6 Acknowledgements

We would like to thank Dr Lesong Conteh and Dr Paula Christen from the London School of Economics (LSE)-International Science Partnership Fund (ISPF) for their helpful review and feedbacks. Collaboration through their project ‘From data to delivery: understanding vaccine and diagnostics markets in Senegal’ has greatly improved this work.

## 8 Appendix

The Appendix provides a detailed overview of the input parameters used in the analysis. Table 4 summarizes epidemiological parameters related to cervical cancer in Senegal, including age-specific incidence and mortality rates, stage distribution, HPV genotype prevalence, disability weights, and five-year survival probabilities. Table 5 presents the unit costs of cervical cancer treatment from the government perspective, disaggregated by disease stage. Table 6 reports HPV vaccine program cost parameters, including vaccine and supply prices, international handling and delivery fees, wastage rates, and incremental health system costs.

